# The associations of chronotype and shift work with rheumatoid arthritis

**DOI:** 10.1101/2022.07.07.22277352

**Authors:** TD Butler, RJ Maidstone, MK Rutter, J McLaughlin, DW Ray, JE Gibbs

## Abstract

**Introduction:** The circadian clock regulates multiple aspects of human physiology including immunity. People have a circadian preference termed chronotype. Those with an evening preference may be better suited to shift work, but also carry higher risk of adverse health. Shift work leads to misalignment of circadian rhythms and is associated with increased risk of inflammatory disease such as asthma and cancer. Here, we investigate the association between chronotype, shift work and rheumatoid arthritis (RA).

**Methods:** The associations between exposures of shift work and chronotype on risk of RA were studied in up to 444,210 UK Biobank participants. Multivariate logistic regression models were adjusted for covariates including age, sex, alcohol intake, smoking history and BMI.

**Results:** After adjusting for covariates, individuals with a morning chronotype had lower odds of having RA (OR 0.93, 95% CI 0.88 to 0.99) when compared to intermediate chronotypes. The association between morning chronotype and lower odds of RA persisted with a more stringent RA case definition (Covariate-adjusted OR 0.89, 95% CI 0.81 to 0.97). Shift workers had a higher odds of RA (OR 1.22, 95% CI 1.1 – 1.36), that attenuated to the null after covariate adjustment.

**Conclusions:** These data implicate circadian rhythms in RA pathogenesis. Further studies are required to determine the mechanisms underlying this association and understand the potential impact of shift work on chronic inflammatory disease and its mediating factors.

**KEY MESSAGES:** *What is already known about this subject?:*

- Chronotype impacts a person’s ability to adapt to shift work, which contributes to circadian misalignment and is associated with adverse health outcomes including asthma, diabetes and cancer

*What does this study add?:*

- People with morning chronotype have a lower likelihood of RA, compared to intermediate chronotypes
- Shift workers have higher likelihood of RA, compared to day workers, but this relationship may be mediated or confounded by factors such as smoking and BMI

*How might this impact on clinical practice or future developments?:*

- Chronotype is easy to assess and could be combined with other factors to determine an individual’s risk of developing RA and influence decisions about undertaking shift work
- Growing evidence that common chronic inflammatory disorders are affected by circadian misalignment generates further need to investigate the impact of chronotype and shift work on immune function

## INTRODUCTION

Rheumatoid arthritis (RA) is an increasingly prevalent autoimmune chronic inflammatory disease primarily affecting the joints and significantly impacting morbidity and premature mortality (*1, 2*). There is diurnal variation in symptoms of RA, with exacerbations of joint pain and stiffness in the morning, aligned to rhythmic expression of pro-inflammatory serum cytokines such as IL-6 (*3*).

Shift work is becoming increasingly common worldwide; with 10-20% of workers in most countries undertaking night shifts (*4*). Circadian misalignment as a result of night shift work impacts on circadian processes supporting healthy immune function (*5, 6*) and population data shows that shift workers have a higher likelihood of conditions with inflammatory components such as asthma (*7*) and Covid-19 (*8*).

24-hour circadian clocks allow humans to distribute physiological processes across the day, using a molecular clock mechanism to keep time and environmental stimuli such as light:dark cycles to set the time. Individuals have their own circadian phase preference, termed chronotype, with an evening chronotype being associated with lower levels of health and performance (*9*).

There are limited data linking circadian misalignment with RA and therefore we aimed to assess how strongly shift work and chronotype are related to the presence of RA in UK Biobank participants. As incidence of shift work and RA increases (*1*), these data could have important public health implications.

## METHODS

Between 2007 and 2010, the UK Biobank recruited 502,540 National Health Service-registered participants aged 40-69 years old (*10*). Baseline assessments conducted at 22 UK study centres collected data on lifestyle, medical history, occupation and work patterns.

### Exposures

a. *Chronotype*: Self-reported chronotype data were collected at baseline in UK biobank participants (n=444,210) by asking the following question that had four answer options: ‘Do you consider yourself to be…’ a) “definitely a ‘morning’ person?”, which we defined as ‘morning chronotypes’; b) “definitely an ‘evening’ person?”, which we defined as ‘evening chronotypes’. An ‘Intermediate chronotype’ group was formed by combining responses: c) “more a ‘morning’ than ‘evening’ person?”; and d) “more an ‘evening’ than ‘morning’ person?”. Individuals answering “do not know” were excluded from analysis (n=4,032).
b. *Shift work*: Shift work analysis was restricted to participants in paid employment or self-employment at baseline (n=286,825, age range 37-72 years). Shift workers were sub-categorised as working permanent night shifts or working irregular shifts, which included shift workers who never, rarely or sometimes worked night shifts.

#### Outcome

Cases of prevalent RA included patients with self-reported, doctor-diagnosed RA (code 20002) or patients with International Classification of Diseases (ICD)-9/ICD-10 codes for RA diagnosis (ICD-10 codes: M05, M06 and ICD-9 code: 7140), which represent diagnoses recorded during secondary care stays prior to the UK Biobank baseline assessment. This identified 5,659 cases (1.3% of all UK Biobank participants), of which 3,537 were included in the chronotype analysis and 2,201 were included in the shift work analysis. Sub-group analysis to increase RA case specificity included patients with self-reported, doctor-diagnosed RA who, in addition reported either a prescribed medication to treat RA (supplementary table 1) or had an ICD code for RA.

### Statistical analysis

Descriptive data are presented as mean (standard deviations (SD)) or percentages. Multivariable-adjusted logistic regression models generated ORs and 95% asymptotic confidence intervals (CI) describing the strength of relationships between shift work and chronotype with prevalent RA. Models were serially adjusted for the following covariates: Model 1: age, sex, ethnicity and Townsend deprivation index; model 2: model 1 covariates plus sleep duration, alcohol intake, smoking status and length of working week; Model 3: model 2 covariates plus BMI (*11*). Analysis was performed using statistical programming software R. p<0.05 was considered statistically significant.

## RESULTS

### Chronotype

The characteristics of participants providing chronotype data are detailed in Table 1. When compared to intermediate chronotypes, morning chronotypes were less likely to be smokers and consumed less alcohol, but had similar BMI. Compared to intermediate chronotypes, evening chronotypes were younger, more likely to be male, smokers, with increased alcohol consumption and a diagnosis of depression.

**Table 1.**
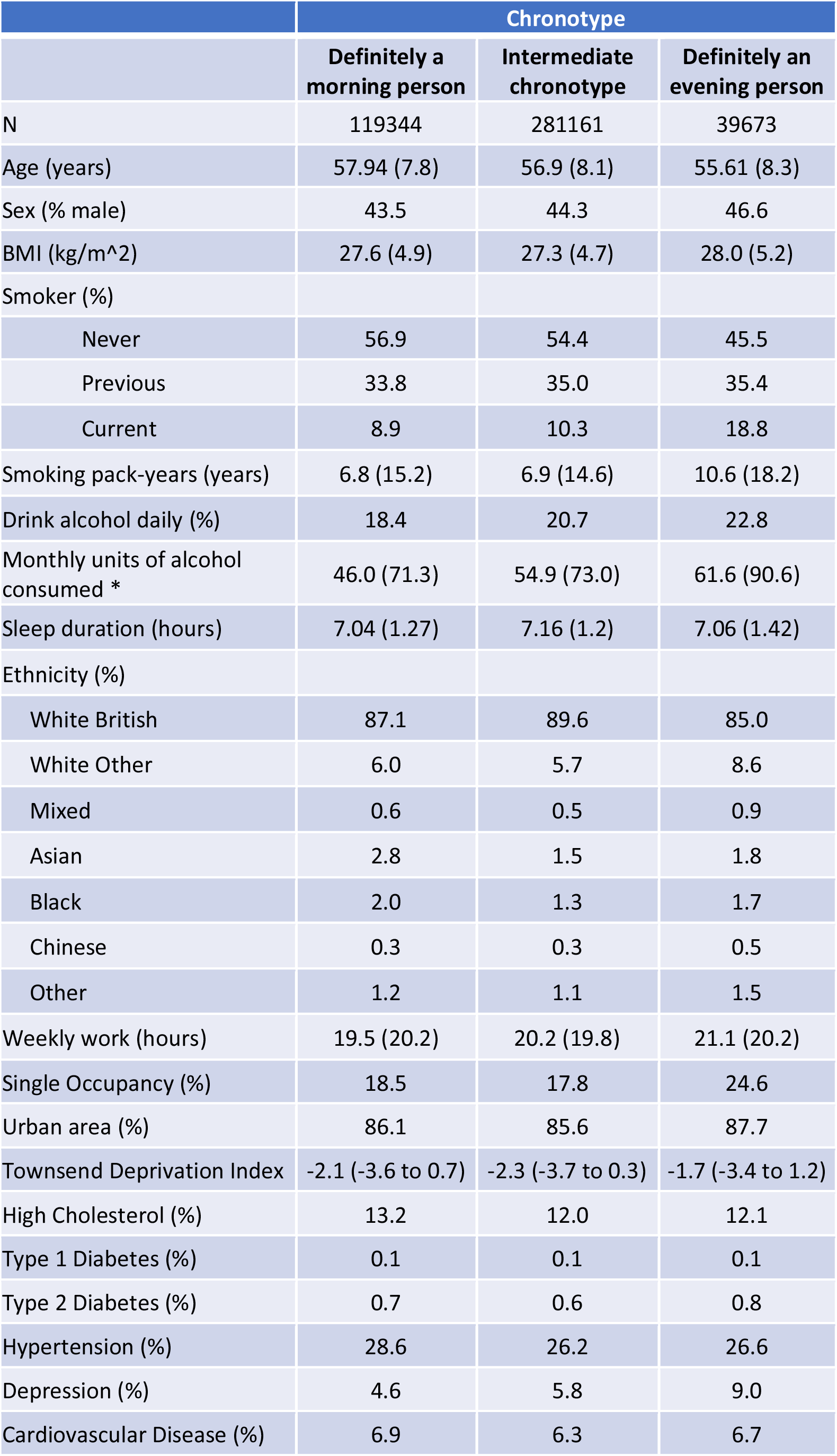
Participant characteristics by chronotype (N = 440,178) Data are mean (SD) or percentages. Townsend Deprivation Index is a measure of socioeconomic deprivation, where higher scores correlate with greater deprivation. Doctor-diagnosed comorbidities including type 1 diabetes, type 2 diabetes, hypertension, depression and cardiovascular disease were recorded by participants on touchscreen questionnaire at UK Biobank enrolment.

UK Biobank participants with an extreme morning chronotype did not have lower odds of RA in model 1 (OR 0.96, 95% CI 0.91 to 1.02), but on further covariate adjustment, demonstrated lower odds of RA in model 2 (OR 0.93, 95% CI 0.88 to 0.99) and model 3 (OR 0.93, 95% CI 0.88 to 0.99), compared to intermediate chronotypes (Figure 1).

**Figure 1.**
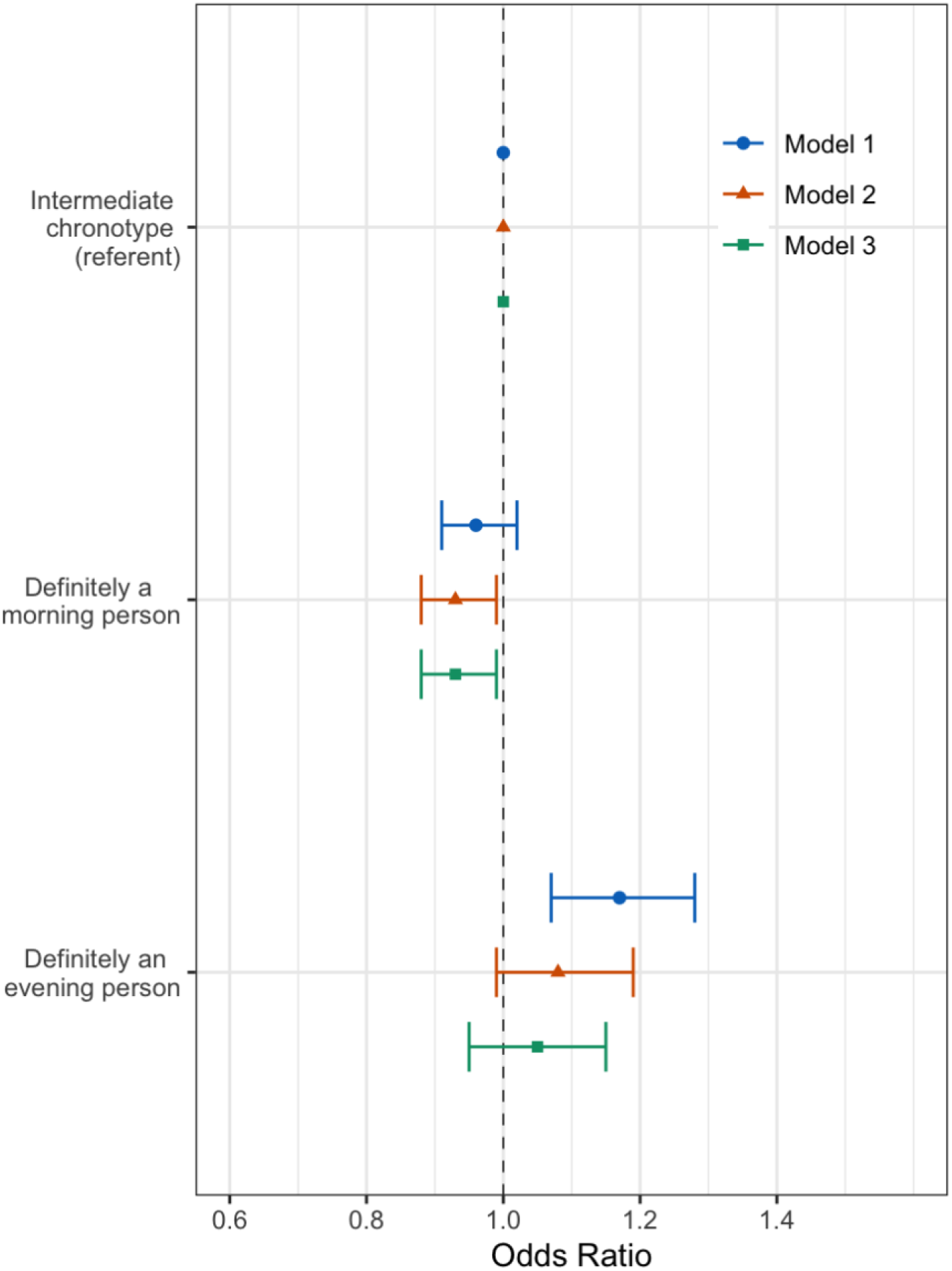
Adjusted odds of rheumatoid arthritis by chronotype (n=440,178) Forest plots of adjusted OR and 95% CIs for RA by chronotype, with intermediate chronotypes as a referent population. Three multivariate logistic regression models were used. Model 1 (circle): age, sex, ethnicity and Townsend Deprivation index; Model 2 (triangle): sleep duration, alcohol intake, smoking status and length of working week, plus model 1 covariates; Model 3 (square): BMI in addition to model 2’s covariates.

In the sub-group with self-reported or doctor-diagnosed RA who, in addition were either prescribed RA medication or had an RA ICD code, significant associations between morning chronotype and a lower likelihood of RA were observed in all three models: model 1 (OR 0.91, 95% CI 0.84 to 0.99), model 2 (OR 0.89, 95% CI 0.81 to 0.97) and model 3 (OR 0.89, 95% CI 0.81 to 0.97) when compared to intermediate chronotypes (Supplementary figure 1).

Extreme evening chronotypes had a higher odds of RA (OR 1.17, 95% CI 1.07 to 1.28) when compared to intermediate chronotypes in the minimally adjusted model (model 1). However, the association became non-significant in model 2 (OR 1.08, 95% CI 0.99 to 1.28) and model 3 (OR 1.05, 95% CI 0.95 to 1.15) (Figure 1).

In the sub-group analysis, associations between evening chronotype and RA were significant in model 1 (OR 1.15, 95% CI 1.01 to 1.30), but not in model 2 (OR 1.05, 95% CI 0.92 to 1.19) or model 3 (OR 1.04, 95% CI 0.91 to 1.18) (Supplementary figure 1).

### Shift work

In participants in paid employment or self-employment, 83% were day-only workers and the remaining 17% worked shifts, of which 51% included night shifts. Compared to day workers, shift workers were more likely to be male, smokers, work longer weeks and drink less alcohol (Table 2). Night shift workers were more likely to be evening chronotypes and work in technical occupations.

**Table 2.** Participant characteristics by current night shift work exposure (N = 286,825) Data are mean (SD) or percentages.

|  | Current work schedule |  |  |  |
| --- | --- | --- | --- | --- |
|  | Day workers | Shift work, but never or rarely night shifts | Irregular shift work including nights | Permanent night shift work |
| N | 236897 | 24560 | 18226 | 7142 |
| Age (years) | 52.9 (7.1) | 52.5 (7.1) | 51.1 (6.9) | 51.5 (6.9) |
| Sex (% male) | 46.6 | 47.5 | 62.4 | 61.4 |
| BMI (kg/m<sup>2</sup>) | 27.1 (4.7) | 27.8 (5.0) | 28.2 (4.9) | 28.5 (4.9) |
| Smoker (%) |  |  |  |  |
| Never | 58.1 | 53.7 | 52.8 | 52.0 |
| Previous | 31.9 | 32.1 | 30.5 | 30.0 |
| Current | 9.8 | 13.9 | 16.2 | 17.7 |
| Smoking pack-years | 20.1 (16.1) | 22.9 (17.5) | 24.3 (17.8) | 25.7 (18.4) |
| Daily alcohol intake (%) | 20.5 | 16.9 | 16.0 | 10.2 |
| Sleep Duration (h) | 7.1 (1.0) | 7.0 (1.2) | 6.9 (1.3) | 6.7 (1.5) |
| Morning Chronotype (%) | 23.3 | 25.5 | 22.9 | 19.2 |
| Evening Chronotype (%) | 8.0 | 7.9 | 9.8 | 16.9 |
| Ethnicity |  |  |  |  |
| White British | 88.5 | 83.3 | 79.9 | 81.0 |
| White Other | 6.5 | 7.1 | 7.0 | 6.0 |
| Mixed | 0.7 | 0.9 | 1.0 | 0.9 |
| Asian | 1.7 | 3.6 | 3.8 | 3.4 |
| Black | 1.4 | 2.7 | 4.9 | 5.5 |
| Chinese | 0.3 | 0.5 | 0.5 | 0.7 |
| Other | 0.1 | 0.1 | 0.1 | 0.1 |
| Weekly work hours | 34.2 (13.2) | 35.0 (13.2) | 39.3 (14.6) | 39.6 (13.7) |
| Single Occupancy (%) | 15.6 | 18.8 | 18.7 | 18.4 |
| Urban area (%) | 86.0 | 89.6 | 89.3 | 91.0 |
| Townsend Index | -2.2 (-3.7 to 0.2) | -1.3 (-3.2 to 1.6) | -1.2 (-3.2 to 1.8) | -1.0 (-3.0 to 2.1) |
| High Cholesterol (%) | 7.9 | 8.6 | 8.5 | 9.3 |
| All Diabetes (%) | 3.2 | 4.3 | 4.6 | 4.7 |
| Type I Diabetes (%) | 0.1 | 0.1 | 0.1 | 0.1 |
| Type II Diabetes (%) | 0.4 | 0.6 | 0.6 | 0.6 |
| Hypertension (%) | 20.3 | 22.2 | 22.1 | 23.3 |
| Depression (%) | 4.6 | 5.5 | 4.5 | 4.8 |

In model 1, people undertaking any shift work had higher odds of having RA (OR 1.22, 95% CI 1.1 to 1.36) (Figure 2). Further covariate adjustment in model 2 attenuated this association (OR 1.12, 95% CI 1.0 to 1.25), p<0.0001), with attenuation to the null in model 3 (OR 1.1, 95% CI 0.98 to 1.22).

**Figure 2.**
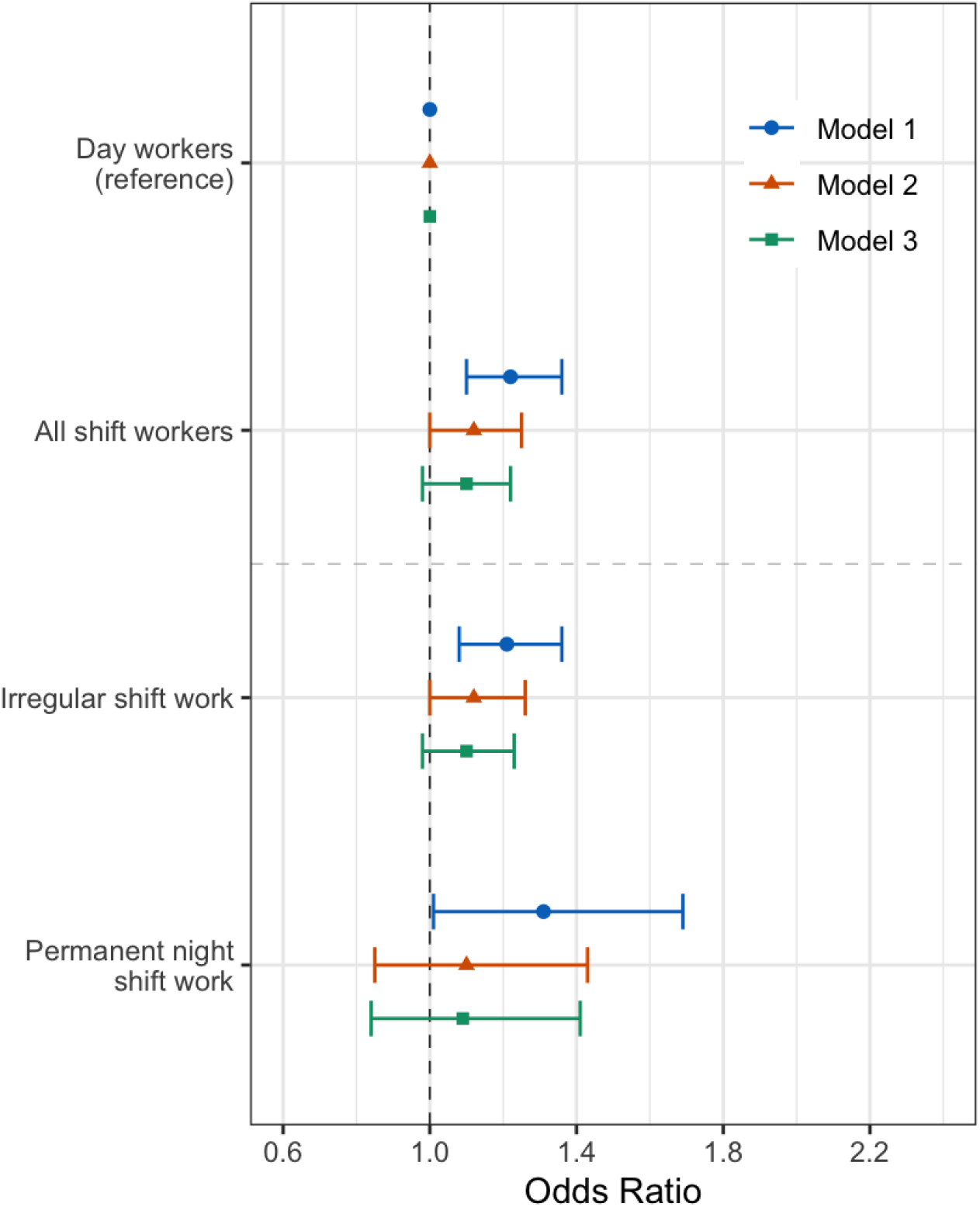
Adjusted odds of rheumatoid arthritis in shift workers (n=286,825) Forest plots of adjusted OR and 95% CIs for RA in all shift workers and shift workers stratified by current shift pattern, with day workers as a referent population. Three multivariate logistic regression models were used. Model 1 (circle): age, sex, ethnicity and Townsend Deprivation index; Model 2 (triangle): sleep duration, alcohol intake, smoking status and length of working week, plus model 1 covariates; Model 3 (square): BMI in addition to model 2’s covariates.

Irregular shift patterns (OR 1.21, 95% CI 1.08 to 1.36) and permanent night shifts (OR 1.31, 95% CI 1.01 to 1.69), were both linked to a higher likelihood of RA, when compared to day workers in model 1 (Figure 2). However, for irregular shift workers, further adjustment for covariates in model 2 (OR 1.12, 95% CI 1.0 to 1.26) and model 3 (OR 1.1, 95% CI 0.98 to 1.23) attenuated the association. Similarly, the odds of having RA in permanent night shift workers was attenuated to the null in model 2 (OR 1.1, 95% CI 0.85 to 1.43) and 3 (OR 1.09, 95% CI 0.84 to 1.41).

In the sub-group analyses, irregular shift work (143 cases) and permanent night shift work (25 cases) were not significantly related to prevalent RA (Supplementary table 2).

### Interaction between shift work and chronotype

To understand the influence of chronotype on the association between shift work and rheumatoid arthritis, permanent night shift workers were stratified by chronotype. There were no significant associations between permanent shift workers and rheumatoid arthritis, stratified by chronotype and no significant interaction by chronotype status (P_interaction_ = 0.16) (Supplementary table 3).

## DISCUSSION

This study analysed data from over 440,000 UK Biobank participants and demonstrates lower adjusted odds of RA in individuals with a morning chronotype, which persists after covariate adjustment when a more stringent RA case selection is used. In addition, this study demonstrates an association between shift work and prevalent RA which is attenuated on covariate adjustment.

Our results are contrary to a recent paper, where authors used the Munich Chronotype Questionnaire to report an earlier mid-point of sleep in 121 RA cases from the Netherlands, compared to healthy controls (*12*). Our study involved a much larger sample size and adjustment for multiple covariates, but our case definition specificity may be lower due to reliance on a combination of doctor-diagnosed RA and ICD codes, rather than rheumatologist-diagnosed RA. In our analysis, morning, compared to intermediate chronotype is linked to risk factors associated with higher RA prevalence such as lower daily alcohol intake, shorter sleep duration and shorter working week, and therefore when these factors are accounted for in model 2, the odds of RA associated with morning chronotype is lowered and the association becomes statistically significant. This pattern persists in model 3. Further work is required to understand whether patients with RA have a chronotype shaped by rhythmicity in symptoms, against a morning preference when joints are at their stiffest.

Our study demonstrated higher adjusted odds of RA in shift workers, including those who never or rarely work night shifts and permanent shift workers, when compared to day workers. However, this association attenuated to the null in models 2 and 3. These data would be in keeping with sleep duration, smoking and/or BMI being confounders and/or mediators of the relationship between shift work and RA. Further work will be required to elucidate these potential mechanistic links. Shift workers constantly challenge entrainment of their circadian rhythms and the systemic consequences of desynchrony between central and peripheral circadian oscillators designates shift work a risk factor for chronic diseases such as asthma, diabetes and cancer (*7, 13, 14*). Our data provides evidence that RA should be considered in this list.

This study builds on two previous smaller studies analysing shift work and RA. In a Finnish population of public sector workers, women working night shifts had a higher risk of incident RA when compared to day workers (*15*). In a Swedish case-control study, rotating shift work was associated with a higher risk of incident seropositive RA, whereas permanent night shift work conferred a protective effect (*16*).

A strength of our study is the large population of RA cases from the UK Biobank, which also contains data to enable robust adjustment for confounders that impact chronotype, such as age, sex and ethnicity. (*17*) The protective association between morningness and RA persisted through these models. In patients with RA, inflammatory cytokines peak in the morning (*3*). Taken in context of our findings, this raises the interesting possibility that biological clocks in morning types are better aligned to regulate and control inflammatory processes that drive RA pathogenesis, thus avoiding RA development. Further profiling of morning types, including circadian-driven meal timing, activity profiles and sleep hygiene may identify protective factors that could help reduce RA risk in non-morning types and highlight potential chronotherapeutic avenues such as chronotype-driven drug timing. In our study, the trend towards higher odds of RA in evening types did not reach statistical significance. Evening chronotype has been associated with increased severity of chronic inflammatory disease (*18, 19*) and whilst it was not possible to risk stratify RA cases by severity in this study, further work should explore the association of chronotype and RA severity.

The UK Biobank has inherent limitations, for example the participation rate was 5% and skewed towards more healthy individuals. Over half of RA cases had no recorded medication for RA, which raises the possibility of misdiagnosis. To investigate this, sub-group analysis aimed to increase specificity of RA case definition by cross-referencing self-reported RA cases with the presence of either an ICD-10 code for RA or presence of RA medication. Whilst this decreased case numbers by 2,832, the lower odds of RA in extreme morning chronotypes persisted. This gives support to the main case definition and the main findings. In addition, RA case numbers in shift work analyses were low, with only 62 permanent night shift workers identified, impacting the power of results. This may be due to the high levels of withdrawal from paid employment seen in RA patients (*20*).

## CONCLUSION

This study reports an association between morning chronotype and lower risk of RA as well as an increased adjusted odds of RA in shift workers. This adds to the mounting evidence that circadian rhythms are a key factor in RA and adds strength to the idea that misalignment of internal clock phase with the external 24h environment may be important in RA development. Further studies are required to explore the mechanistic links between circadian misalignment and development, or propagation of inflammation seen in RA.

## Data Availability

All data produced in the present work are contained in the manuscript

---

**Supplementary table 1.**
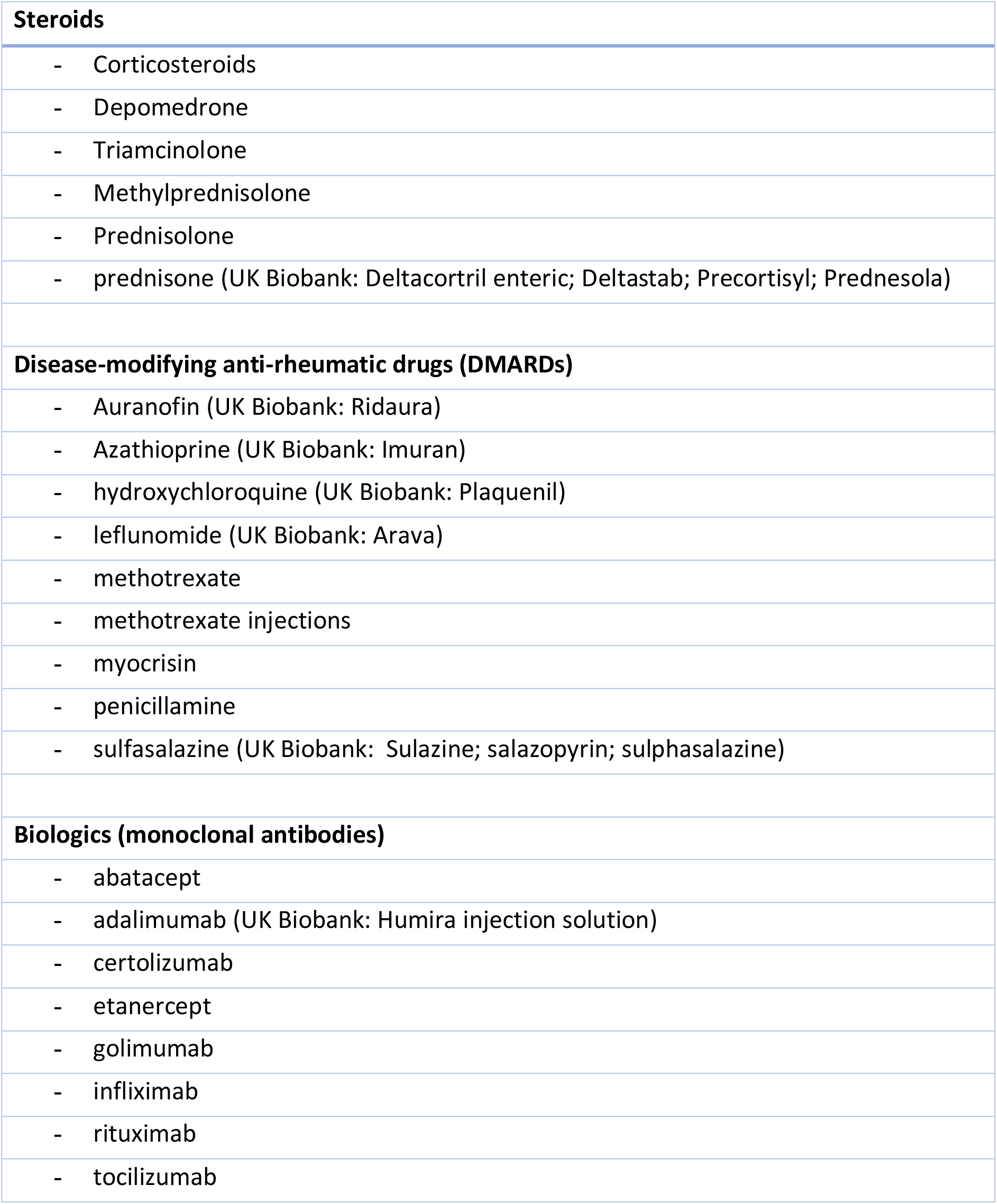
Medications for RA recorded by UK Biobank participants.

|  |
| --- |
| **Steroids** |
| - Corticosteroids |
| - Depomedrone |
| - Triamcinolone |
| - Methylprednisolone |
| - Prednisolone |
| - prednisone (UK Biobank: Deltacortril enteric; Deltastab; Precortisyl; Prednesola) |
| **Disease-modifying anti-rheumatic drugs (DMARDs)** |
| - Auranofin (UK Biobank: Ridaura) |
| - Azathioprine (UK Biobank: Imuran) |
| - hydroxychloroquine (UK Biobank: Plaquenil) |
| - leflunomide (UK Biobank: Arava) |
| - methotrexate |
| - methotrexate injections |
| - myocrisin |
| - penicillamine |
| - sulfasalazine (UK Biobank: Sulazine; salazopyrin; sulphasalazine) |
| **Biologics (monoclonal antibodies)** |
| - abatacept |
| - adalimumab (UK Biobank: Humira injection solution) |
| - certolizumab |
| - etanercept |
| - golimumab |
| - infliximab |
| - rituximab |
| - tocilizumab |

**Supplementary table 2.** Subgroup analysis of adjusted odds (95% CI) of rheumatoid arthritis (self-reported with either medication or ICD code) in shift workers by shift work schedule (N = 282,303). Model 1: age, sex, ethnicity and Townsend Deprivation index; Model 2: sleep duration, alcohol intake, smoking status and length of working week, plus model 1 covariates; Model 3: BMI in addition to model 2’s covariates.

|  | Current work schedule |  |  |
| --- | --- | --- | --- |
|  | Day workers | Irregular shift work | Always |
| **Total cases (% of total sample size)** | 892 (0.38%) | 143 (0.34%) | 25 (0.35%) |
| **Total sample size** | 233,423 | 41,826 | 7,054 |
| **Model 1** | 1 | 0.99 (0.83-1.18) | 1.11 (0.75-1.66) |
| **Model 2** | 1 | 0.91 (0.76-1.09) | 0.94 (0.63-1.4) |
| **Model 3** | 1 | 0.91 (0.76-1.09) | 0.94 (0.63-1.41) |

**Supplementary Table 3.**
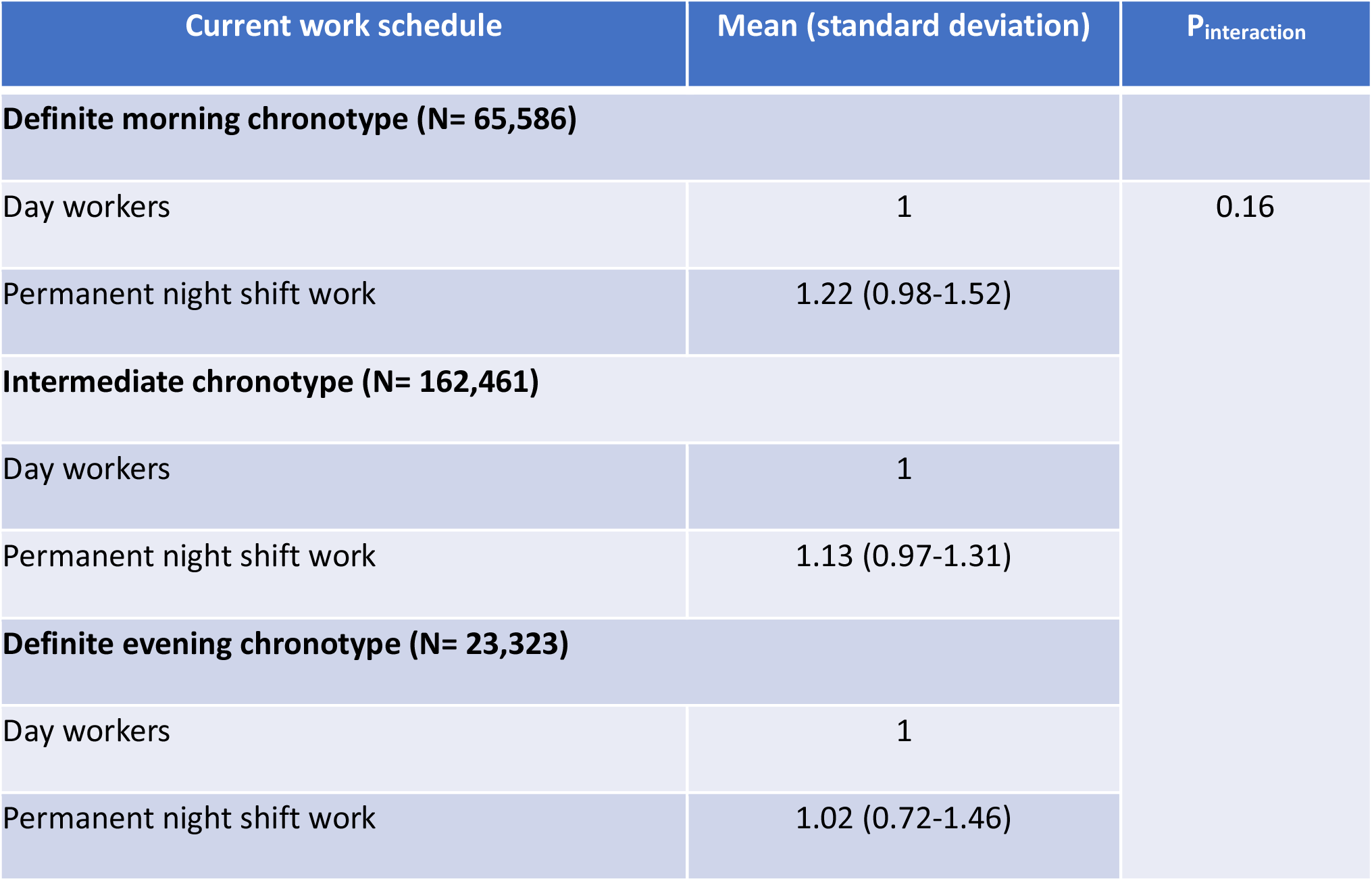
Adjusted odds (95% CI) of rheumatoid arthritis (ICD code or self-reported) by shift work schedule, stratified by chronotype.

**Supplementary Figure 1.**
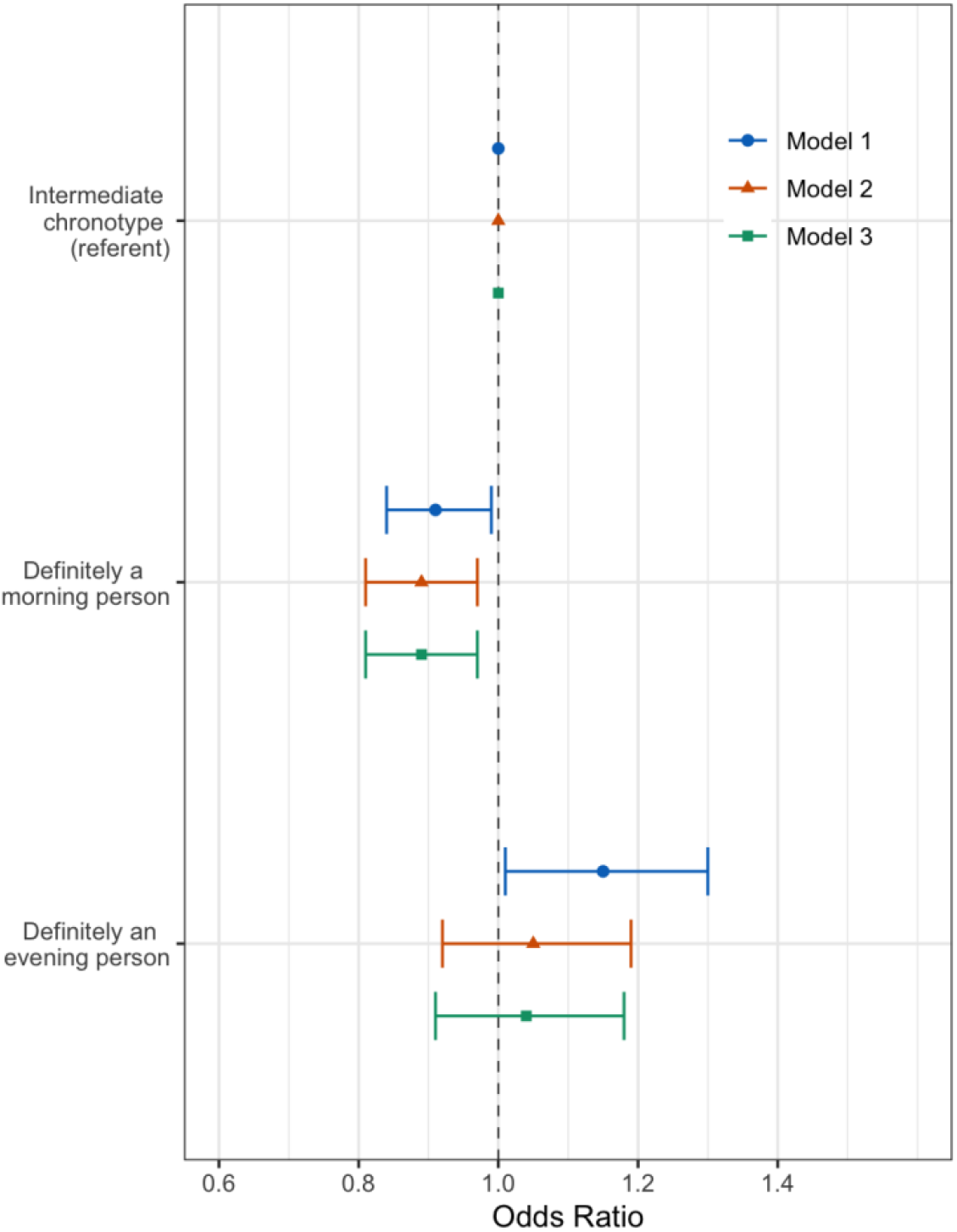
Subgroup analysis of adjusted odds of rheumatoid arthritis (self-reported with either medication or ICD code) by chronotype (n=440,178) Forest plots of adjusted OR and 95% CIs for RA by chronotype, with intermediate chronotypes as a referent population. Three multivariate logistic regression models were used. Model 1 (circle): age, sex, ethnicity and Townsend Deprivation index; Model 2 (triangle): sleep duration, alcohol intake, smoking status and length of working week, plus model 1 covariates; Model 3 (square): BMI in addition to model 2’s covariates.

